# Acute tubulointerstitial nephritis with or without uveitis: a novel form of post-acute COVID-19 syndrome in children

**DOI:** 10.1101/2023.01.23.23284848

**Authors:** Marina Avramescu, Pierre Isnard, Sarah Temmam, Agnès Chevalier, Paul Bastard, Mikael Attia, Romain Berthaud, Marc Fila, Claire Dossier, Julien Hogan, Tim Ulinski, Damia Leguevaques, Férielle Louillet, Edouard Martinez Casado, Jean-Michel Halimi, Sylvie Cloarec, Ariane Zaloszyc, Camille Faudeux, Caroline Rousset-Rouvière, Stéphanie Clavé, Jérôme Harambat, Edouard Rollot, Thomas Simon, Megan Nallet-Amate, Bruno Ranchin, Justine Bacchetta, Florence Porcheret, Josselin Bernard, Amélie Ryckewaert, Anne Jamet, Jacques Fourgeaud, Nicolas Da Rocha, Philippe Pérot, Nicolas Kuperwasser, Naïm Bouazza, Marion Rabant, Jean-Paul Duong Van Huyen, Matthieu P Robert, Julien Zuber, Jean-Laurent Casanova, Marc Eloit, Isabelle Sermet-Gaudelus, Olivia Boyer

## Abstract

**Background:** COVID-19 is a complex multisystem disease, frequently associated with kidney injury. Since the beginning of the COVID-19 pandemic, we observed a striking increase in the incidence of acute tubulointerstitial nephritis (aTIN) without or with uveitis (TINUs) among children. This prompted us to examine whether SARS-CoV-2 might be the underlying trigger.

**Methods:** We conducted a French nationwide retrospective cohort study. We included all consecutive children diagnosed with aTIN or TINUs of undetermined cause between April-2020 and March-2021. SARS-CoV-2 antibody responses were tested by a luciferase immunoprecipitation system and compared to age-matched controls. Immunohistochemistry, immunofluorescence and molecular microbiology analyses were performed on kidney biopsies.

**Results:** Forty-eight children were included with a median age at diagnosis of 14.7 years (9.4-17.6). aTIN and TINUs incidence rates increased 3-fold and 12-fold, respectively, compared to pre-pandemic years. All patients had impaired kidney function with a median eGFR of 31.9 ml/min/1.73m² at diagnosis. Kidney biopsies showed lesions of acute tubulointerstitial nephritis and 25% of patients had fibrosis. No patient had concomitant acute COVID-19. All 16 children tested had high anti-N IgG titers and one had anti-S IgGs. Next-generation sequencing failed to detect any infectious agents in kidney biopsies. However, SARS-CoV-2 RNA was detected by PCR in two kidney samples supporting a potential direct link between SARS-CoV-2 and aTIN/TINUs.

**Conclusions:** We describe a novel form of post-acute COVID-19 syndrome in children, unique in its exclusive kidney and eye involvement, and its distinctive anti-SARS-CoV-2 N+/S-serological profile. Our results support a causal association linking SARS-CoV-2 infection to this newly-reported burst of renal/eye inflammation.

## INTRODUCTION

Multi-organ sequelae of COVID-19 beyond the acute phase of infection are increasingly described as clinical experience expands. In children, acute COVID-19 appears to be generally asymptomatic or mild. Yet, the multisystem inflammatory syndrome in children (MIS-C) may be a severe post-infectious complication following exposure to SARS-CoV-2^1^. During the first pandemic year, we observed a striking increase in the incidence of acute tubulointerstitial nephritis (aTIN) without or with uveitis (TINUs) among children. Causes of aTIN include drugs, infections, and systemic diseases, but often remain undetermined. The rare TINU syndrome associating aTIN and uveitis is considered to result from a still ill-characterized immune-mediated process. The observed increased incidence of idiopathic aTIN/TINUs prompted us to examine whether SARS-CoV-2 might be the initial trigger.

## RESULTS

### Increased incidence of aTIN and TINUs during the pandemic

Between 01/04/2020-31/03/2021, 48 children with a median age of 14.7 years (9.4-17.6) were diagnosed with aTIN (n=25) or TINUs (n=23) of undetermined cause in France (Figure 1a-b) compared to the 8-10 cases/year of idiopathic aTIN recorded in 2018-19, and 46 TINUs cases over an 18-year period between 2000-2018^2^. This translates into a 3-fold and 12-fold increase in the incidence of aTIN and TINUs, respectively (from 1 to 3 per million children, and 0.12 to 1.4 per million children, respectively).

**Figure 1.**
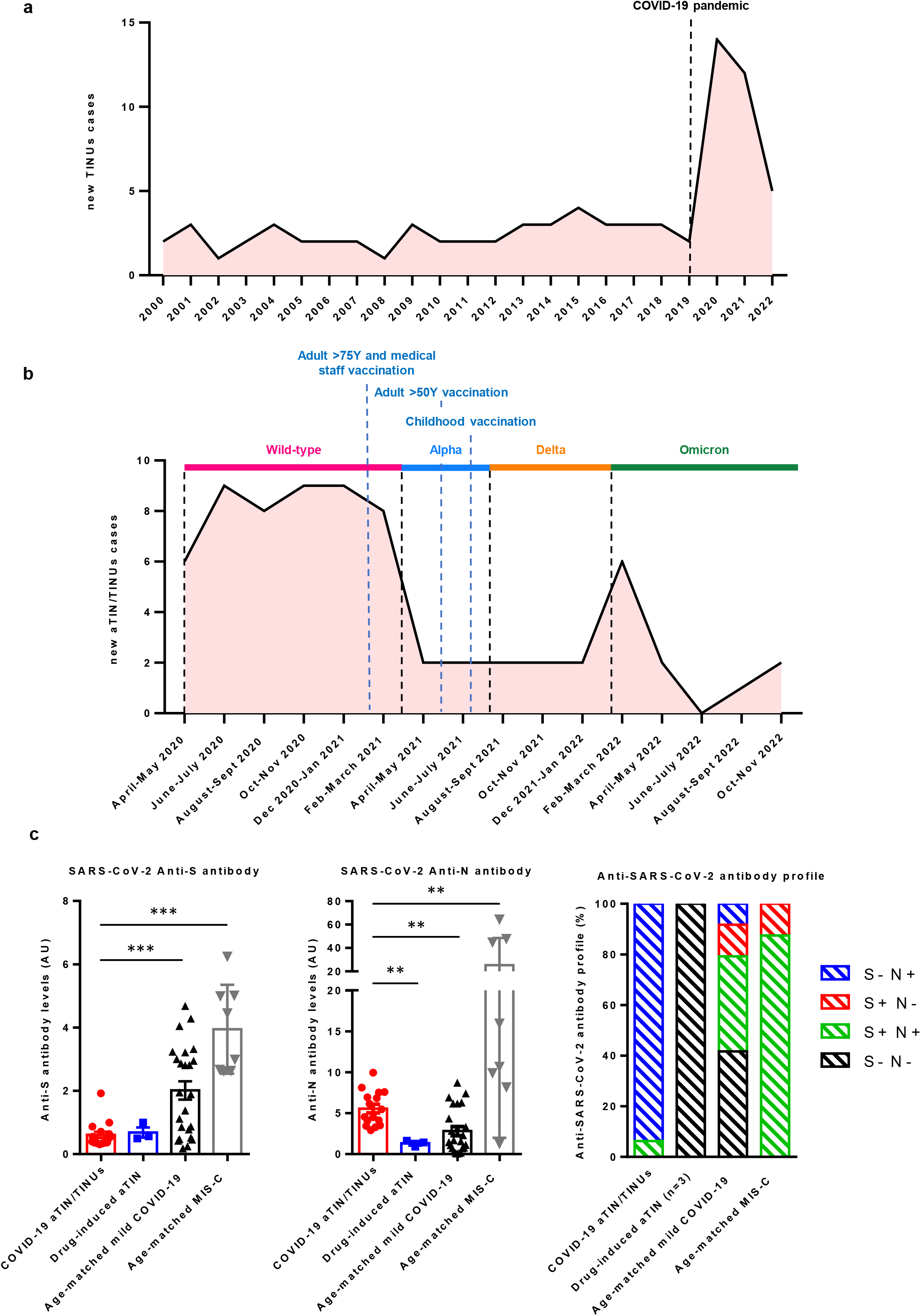
Incidence of nationwide pediatric aTIN/TINUs cases in France and anti-SARS-CoV-2 antibody levels and profiles in children with aTIN/TINUs. **a**. Annual incidence of nationwide pediatric TINUs cases in France from 2000-2022. **b**. Bimonthly incidence of nationwide pediatric aTIN/TINUs cases in France from April 2020 to November 2022. The approximate dates of circulation of the different strains of SARS-CoV-2 and the dates of launch of the vaccination campaigns according to age are shown in the figure. All analyzed cases of aTIN/TINUs were recorded during the circulation of the original virus and the Alpha variant. Interestingly, only 21 cases of aTIN/TINUs were later recorded between April 2021 and November 2022, when the Delta and the Omicron variants were the predominant viral strains. Of note, none of the patients had received any vaccination against SARS-CoV-2 before the diagnosis of aTIN/TINUs as the campaign for children above 12 years was launched on June 15th 2021 in France. **c**. Anti-SARS-CoV-2 anti-S and anti N antibody levels and profiles in children with aTIN/TINUs compared to contemporaneous age-matched children with Ibuprofen-induced aTIN (n=3) or hospitalized for mild COVID-19 (n=24) or MIS-C (n=8), all sampled during the first pandemic year^4^. Wilcoxon-Mann-Whitney U test was applied to test the significance of the difference. ** P < 0.01, *** P < 0.001.

### Virological findings

The SARS-CoV-2 nasopharyngeal swab test was negative in all children tested at diagnosis of aTIN/TINUs (37/48) and only 4/48 had presented symptoms of COVID-19 in the preceding weeks. Routine serological SARS-CoV-2 testing targeting the nucleocapsid (N) protein was negative in all 29 tested children and anti-spike (S) serology was positive only in Patient 9 at onset of the kidney disease and before initiation of therapy, at the local centers (Supplementary Table 1).

In order to investigate the potential association between aTIN/TINUs and COVID-19, we retrospectively performed SARS-CoV-2 serologies using a luciferase immunoprecipitation assay (LIPS) presenting a specificity of 100% and sensitivities ranging from 83.1% (LIPS-N) to 99.4% (LIPS-full S)^3,4^ (Supplementary Material). Serum at the time of the aTIN/TINU diagnosis was available for 16 children. Strikingly, all sera tested positive for high-level anti-N IgGs, whereas only one (Patient 9) had positive anti-S IgGs (Figure 1c, Supplementary Table 1). As controls, three children diagnosed with ibuprofen-induced aTIN during the study period had negative LIPS. Moreover, COVID-19 ELISpot was positive in the five children for whom peripheral blood cells were available. Together, these data were consistent with a SARS-CoV-2 specific immune response resulting from an infection since the patients had not yet been vaccinated against COVID-19, and the available vaccines exclusively elicit an anti-Spike antibody response. We also performed a pseudo-neutralization assay to detect the presence of virus-neutralizing antibodies in 16 samples. Neutralizing activity against SARS-CoV-2 was only detected in two cases, in line with the absence of anti-S antibodies.

We next compared the antibody titers and serologic profiles of patients in this study with those of age-matched controls (9-17 years) hospitalized with mild COVID-19 or MIS-C during the same period (Figure 1c)^4^. Children with MIS-C displayed predominantly the N+/S+ profile, with significantly higher antibody levels than the other two groups. Children with mild COVID-19 had higher levels of anti-S antibodies but lower levels of anti-N antibodies than children with aTIN/TINUs.

### Clinico-biological features

We investigated whether the phenotype of the TINUs observed during the pandemic differed from those of the historical French pediatric cohort of TINUs from 2000-2018 (Supplementary Table 2)^2^. No statistically significant difference was observed. The typical presentation was sudden weight loss, polyuria, fever, and biological markers of inflammation. All patients had reduced estimated glomerular filtration rate (eGFR) at diagnosis (median 31.9 ml/min/1.73m² (8.4-87.8)) leading to kidney biopsy. eGFR rose to 86.0 ml/min/1.73m² (66.8-134.5) at 12-months follow-up, while 32% patients had chronic kidney disease.

### Pathology findings

Thirty-nine (81%) patients underwent a kidney biopsy. Biopsies from the 16 patients with extensive serological work-up were blindly reassessed by three kidney pathologists (Supplementary Table 1). Light microscopy showed acute interstitial nephritis characterized by an infiltrate of predominantly mononuclear cells associated with numerous tubulitis lesions and no immunoglobulin deposits, as classically observed in TINUs (Figure 2). We did not notice significant morphological differences between our cohort and a series of 14 pre-pandemic TINUs except a tendency toward more fibrosis and fewer eosinophils in the present series (Supplementary Table 3). To determine whether this renal inflammation could be directly related to a specific pathogen, we performed metagenomic next-generation sequencing (mNGS) from 11 kidney biopsy samples, and no definite pathogens were identified (Supplementary Table 1).We next performed SARS-CoV-2 RT-PCR and detected low viral loads of SARS-CoV-2 mRNA in 2/11 kidney samples.

**Figure 2.**
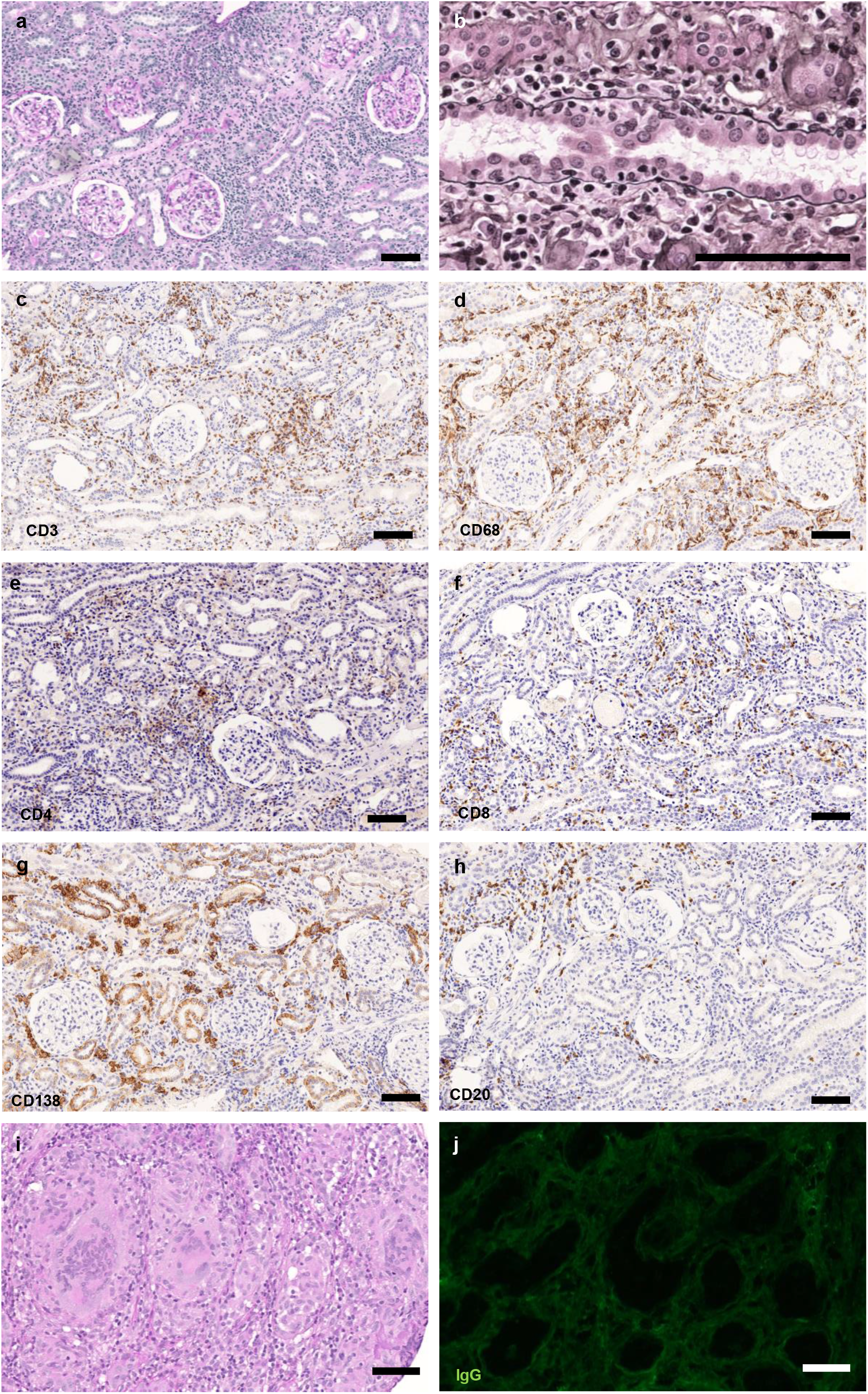
Light microscopy and immunohistochemistry analysis of kidney biopsies. Representative images of optical microscopy. **a**. Light microscopy (x100) using Periodic Acid Schiff staining, showing diffuse interstitial infiltrate mainly composed of mononuclear cells. **b**. Light microscopy (x400) using silver staining showing numerous tubulitis lesions. **c**. Immunohistochemistry analysis (x100) targeting CD3 showing an important T-cell infiltrate. **d**. Immunohistochemistry analysis (x100) targeting CD68 showing an important macrophage infiltrate. **e** and **f**. Immunohistochemistry analysis (x100) targeting CD4 and CD8 showing a relatively balanced distribution of CD8 and CD4 T-cell **g**. Immunohistochemistry analysis (x100) targeting CD138 showing numerous plasma-cells. **h**. Immunohistochemistry analysis (x100) targeting CD20 showing a small B-cell infiltrate. Scale Bar = 100µm. **i**. Light microscopy (x100) using Periodic Acid Schiff staining, showing granulomas with multinucleated giant cells in a minority of patients. **j**. Immunofluorescence analysis (x100) targeting IgG showing no tubular basal membrane deposits.

## DISCUSSION

To our knowledge, this is the first study showing an increased incidence of pediatric aTIN and TINUs superimposed to the first wave of the COVID-19 pandemic. To date, the association of aTIN and COVID-19 has been suggested in few cases^5,6^. We inferred from our study that aTIN/TINUs could be considered as a novel form of post-COVID-19 disease in children. As a matter of fact, epidemiological data, positive SARS-CoV-2 serologies and ELISpot in all tested patients (16/16 and 5/5, respectively), and *in situ* detection of SARS-CoV-2 in two kidney biopsies, strongly support a causal link. COVID-19 was asymptomatic in 44/48 (90%) children at the acute phase, as reported in most post-COVID-19 MIS-C series^1^.

Notably, patients had a unique N+/S-serological profile, which differs from those observed in adults with COVID-19 and children with MIS-C who develop both anti-S and anti-N IgGs. Similarly, children with mild COVID-19 predominantly generate anti-S IgGs^4^. The presence of anti-SARS-CoV-2 IgGs in all tested children in the present series further exemplifies the post-infectious nature of COVID-19-related aTIN/TINUs. Moreover, the lack of detection of SARS-CoV-2 mRNA in most kidney samples further supports a resolved infection. Of interest, the clinical and histological features in the present series were similar to those of pre-pandemic TINUs, and affected mostly adolescents^2^. This suggests that SARS-CoV-2-induced aTIN/TINUs cases share common pathogenic pathways with other forms.

We therefore propose that SARS-CoV-2 could prime autoinflammation through molecular mimicry^7^, as reported in other post-COVID-19 long-term symptoms (Long COVID-19). SARS-CoV-2 uses the receptors angiotensin-converting enzyme II (ACE2) and transmembrane protease serine 2 (TMPRSS2) for cell entry and protein S priming, respectively. Importantly, these two proteins are expressed in renal tubules and in eye components^8,9^. Therefore, we hypothesize that both tissues were infected during the acute phase, triggering an overly potent and long-lasting detrimental immune response, despite subsequent clearance of the virus. Of note, the presence of a predominant lympho-histiocytic infiltrate and the absence of immunoglobulin deposits are in favor of a cell-mediated mechanism.

Overall, our relatively large cohort of this rare disease provides additional insights into the pathophysiology of TINUs and suggests that SARS-CoV-2 should be considered among the infectious agents responsible for pediatric aTIN/TINUs.

The limitations of this study are mainly the recruitment through biopsy-based registry, potentially subject to selection bias, the retrospective design precluding fresh sampling for functional immunological testing, including T-cell response to IFN, and the absence of available serological samples from pre-pandemic aTIN/TINUs patients. However, the three contemporaneous children with ibuprofen-related aTIN/TINUs had a negative SARS-CoV-2 LIPS serology.

Interestingly, the aTIN/TINUs incidence declined after April 2021. The vaccination campaign, herd immunity and emergence of new variants might have contributed to this trend. Nevertheless, our data should raise awareness that post-COVID-19 aTIN/TINUs may be responsible for chronic renal damage in adolescents that may compromise kidney function in adulthood.

## Data Availability

All data produced in the present work are contained in the manuscript

## ACKNOWLEDGMENTS

We are grateful to patients and families for their participation to this study. We are also very grateful to Prof Alain Fischer for his critical review of the manuscript and his wise and relevant advice.

## AUTHOR CONTRIBUTIONS

MAv and PI contributed equally to the work. MAv designed the study, handled regulatory ethics submissions, collected and analyzed clinical and serological data, collected available samples, wrote the first draft of the manuscript. MAv contacted PI to collaborate on this study. PI analyzed the histology data, designed and interpreted microbiological molecular data, wrote the manuscript. ST and AC contributed equally to the work. ST performed and analyzed the serological assays, edited the manuscript. AC collected and analyzed clinical data, edited the manuscript.

PB performed the interferon experiments. MAt performed the pseudoneutralisation assay. AJ and JF performed the NGS on kidney biopsies. NDR and PP assisted with serological testing. NK proof edited the manuscript in English and critically reviewed its scientific content. MRa and JPD analyzed the kidney histology. MRo reviewed the ophthalmological data and examined the patients where possible. JZ and JLC critically reviewed the manuscript with their expertise in immunology. ME led the virological experiments. ISG coordinated the COVID-19 cohort in children locally, handled regulatory ethics submissions, analyzed the data and edited the manuscript. OB designed the study, coordinated all experiments, and wrote the manuscript. All the authors followed the patients, collected clinical data, facilitated the study and declare they have seen and approved the final version of the manuscript.

## SUPPLEMENTARY MATERIAL

### Supplementary Methods

#### Supplementary Results

Supplementary Table 1. Histological, immunohistochemical and virological findings in the 16 COVID-19-associated aTIN/TINUs children with available sera stored at onset of kidney disease.

Supplementary Table 2. Clinical and biological characteristics of the 48 children of the present cohort compared to the historical nationwide French cohort of children diagnosed with TINUs from 2000-2018

Supplementary Table 3. Comparison of histological lesions in the 16 children with COVID-19-associated aTIN/TINUs and available sera, with a series of 14 children with pre-pandemic TINUs.

Supplementary Table 4. Primary antibody list.

Supplementary Figure 1. Seasonal coronaviruses seroprevalences between SARS-COV-2 N+ and N-patients

Supplementary Figure 2. Acute polymorphonuclear leukocytes infiltrate on patient 1 kidney biopsy

Supplementary Figure 3. light microscopy showing interstitial fibrosis and tubular atrophy

Supplementary Figure 4. Immunohistochemical detection of N-protein of SARS-CoV-2

Supplementary Figure 6-5. Neutralizing auto-antibodies (Abs) against IFN-α2, IFN-ω or IFN-β in children with COVID-19-related aTIN/TINUs.

### Supplementary References

