## SUPPLEMENTARY DATA for "Acute tubulointerstitial nephritis with or without uveitis: a novel form of post-acute COVID-19 syndrome in children"

**CONTENT**

**Supplementary Methods**

**Supplementary Results**

Supplementary Table 1. Histological, immunohistochemical and virological findings in the 16 COVID-19-associated aTIN/TINUs children with available sera stored at onset of kidney disease.

Supplementary Table 2. Clinical and biological characteristics of the 48 children of the present cohort compared to the historical nationwide French cohort of children diagnosed with TINUs from 2000-2018

Supplementary Table 3. Comparison of histological lesions in the 16 children with COVID-19-associated aTIN/TINUs and available sera, with a series of 14 children with pre-pandemic TINUs.

Supplementary Table 4. Primary antibody list.

Supplementary Figure 1. Seasonal coronaviruses seroprevalences between SARS-COV-2 N+ and N- patients

Supplementary Figure 2. Acute polymorphonuclear leukocytes infiltrate on Patient 1 kidney biopsy

Supplementary Figure 3. light microscopy showing interstitial fibrosis and tubular atrophy

Supplementary Figure 4. Immunohistochemical detection of N-protein of SARS-CoV-2

Supplementary Figure 5. Neutralizing auto-antibodies (Abs) against IFN-α2, IFN-ω or IFN-β in children with COVID-19-related aTIN/TINUs.

**Supplementary References**

**SUPPLEMENTARY METHODS**

**Study design and patients**

We conducted a nationwide multicenter, retrospective cohort study among all French pediatric nephrology units. All pediatric nephrologists were contacted through the *Société Française de Néphrologie Pédiatrique*. We included all consecutive children (aged 0 to 18 years) diagnosed with aTIN or TINUs of undetermined cause during the first pandemic year (between April 2020 and March 2021) in any of the 16 French Pediatric Nephrology units. We excluded 12 cases with a clearly identified cause of aTIN (toxics, infection, sarcoidosis). Patients included in the study had no identified toxic or medication exposure in the weeks preceding aTIN/TINUs diagnosis.

We also contacted all 32 adult nephrology units in the university hospitals in France via the *Société Francophone de Néphrologie, Dialyse et Transplantation* and we only identified 3 additional patients meeting the inclusion criteria, aged 15 to 17 years, who were included in the study. No adult patients with the same clinico-pathological characteristics were identified.

The total number of national aTIN/TINUs cases from 2019 and 2018 was recorded as well as the total number of national aTIN/TINUs cases between April 2021 and December 2022. Data collection was based on patients' medical records. The estimated glomerular filtration rate (eGFR) was calculated using Schwartz’s 2009 formula. Regarding uveitis diagnosis, the Standardization of Uveitis Nomenclature criteria were used^1^.

**SARS-CoV-2 serological assays**

All serological analyses were performed on sera collected at the time of presentation of the kidney disease, before initiation of therapy, and were conducted locally. For patients with remaining stored serum from the day of diagnosis, serologies were centrally reassessed at the Necker Enfants-Malades Hospital (Abbott serologies) and Pasteur Institute (LIPS serologies). The SARS-CoV-2 IgG II Quant assay (Abbott Diagnostics, Illinois, USA) was performed on the Abbott Alinity platform according to the manufacturer’s instructions. This assay is a chemiluminescent microparticle immunoassay used for the detection of IgG antibodies against the spike receptor-binding domain (RBD) or the nucleocapsid antigen of SARS-CoV-2 in serum specimens. Fifty arbitrary units per milliliter (AU/ml) and above are considered positive. Clinical sensitivity of IgG assay increased proportionally over time following RT-PCR-confirmed date (74% within 10 days and 100% within 20 days) and clinical specificity was 100% in the samples from the pre-COVID-19 period^2^.

We used LIPS to detect serum antibody responses to SARS-CoV-2 spike (S) and nucleocapsid (N) proteins from stored sera that were obtained at the day of diagnosis, where available. LIPS was performed as previously described^3,4^. Recombinant antigens were designed as follows: SARS-CoV-2 spike domains S1 (residues 1-698), S2 (residues 686-1208) and RBD (residues 319-336), along with the full S ectodomain (residues 1-1208) and the carboxy-terminal region of the nucleoprotein (residues 233-419) were expressed in fusion with NanoLuciferase in human HEK-293 cells (Thermo Fisher Scientific). Antigens were recovered directly from crude cell lysates (nucleoprotein-based constructs) or from the supernatant (spike-based constructs) without purification, and quantified on a Centro XS3 LB 960 luminometer (Berthold Technologies, France) by adding the luciferase substrate. Patients’ sera were incubated for one hour with each of the five antigens by adding 10 µL of serum to 108 RLU (relative luminescence unit) of antigen; then the immune complexes were precipitated onto a filter plate with protein A/G-coated beads for one hour. After washing, the luminescence was measured with the luminometer. The signal-to-noise ratio, proportional to the initial antibody titer was calculated. Sensitivity and specificity were previously evaluated^4–7^. Moreover, to further confirm the specificity of our SARS-CoV-2 anti-N LIPS assay, we determined anti-N positivity of the four seasonal coronaviruses (HKU1, OC43, 229E, NL63) in our SARS-CoV-2 N+ patients compared to our SARS-CoV-2 N- patients from the Eurosurveillance study^3^. Interestingly, all four seasonal coronaviruses seroprevalences were comparable between SARS-CoV-2 N+ and N- patients, further emphasizing the specificity of our SARS-CoV-2 anti-N LIPS assay (Supplementary Figure 1).

We next compared the serological profiles of with aTIN/TINUs to that of children with Ibuprofen-induced aTIN (n=3) or hospitalized for mild COVID-19 (n=24) or MIS-C (n=8), all sampled during the first pandemic year^3^.

**SARS-CoV-2 neutralizing assay**

Sera were mixed and co-incubated with pseudotyped vector, enveloped with spike protein from Wuhan, at room temperature for 30 min^8^. The mixture was then plated on white tissue culture 96-well plates (Costar) with 20,000 HEK 293T-hACE2 cells (ATCC) per well in suspension in culture medium [Dulbecco’s modified Eagle’s medium–no phenol red (Gibco), 10% fetal calf serum (Gibco), 1% penicillin/streptomycin (Gibco) and 2mM L-Glutamine (Gibco)]. To prepare the cell suspension, cell flasks were washed with Dulbecco’s PBS (DPBS) (Gibco), and a single-cell suspension was made in DPBS + 0.1% EDTA (Promega) to preserve the integrity of the hACE2 protein. After 48 hours, Bright-Glo Luciferase (Promega) was added to the cells (1:1) and bioluminescence was measured using the Centro XS3 LB 960.

A cohort of negative sera was used to determine the threshold. To calculate the percentage of neutralization, the following equation was used: (1 - (measured value/threshold)) X 100.

**Histological analysis**

Histopathological examinations (optical microscopy and immunofluorescence) were performed by pathologists in each center. Sixteen kidney biopsies were then centralized and blindly reassessed by three kidney pathologists. Briefly, kidney biopsies were fixed in formalin, alcohol and acetic acid and paraffin embedded. Four-µm sections were stained with hematoxylin-eosin, periodic acid–Schiff, Masson trichrome, and methenamine silver. Lesion quantifications were made according to the Banff Classification of Renal Allograft Pathology (2018) for interstitial inflammation, tubulitis and interstitial fibrosis.

**Immunohistochemistry and Immunofluorescence**

Immunohistochemical (IHC) characterization and tubular immunofluorescence (IF) were performed on a panel of 11 kidney biopsies. For IHC, an automated BOND-III (Leica Biosystems) stainer was used. Briefly, 4-µm sections of paraffin-embedded kidneys were submitted for appropriate antigen retrieval. Sections were then incubated with different antibodies. Antibody name, reference, dilution and final concentration are provided in the Supplementary Table 4**.** The degree of immune cell infiltrate was blindly determined on IHC sections using a semi-quantitative score: absence (0-10%), mild (10-25%, 1), moderate (25-50%, 2) and severe (50%, 3). For SARS-CoV-2 anti-N antibody validation (Novusbio), IHC was performed on formalin-fixed paraffin-embedded samples of control and SARS-CoV-2 infected Vero cells (Gift from Giovanna Barba-Spaeth, Structural Virology Unit, Institut Pasteur) prepared using the Cytoblock Cell Block Preparation System (Thermo Scientific) according to the manufacturer’s instructions. Immunofluorescence was performed on frozen kidney biopsies with an antibody targeting the heavy chains of IgG using the automated BOND-III.

**Metatranscriptomics**

Molecular analyses were performed on a panel of 11 kidney biopsies. Nucleic acids from nitrogen-frozen and Optimal Cutting Temperature-embedded kidney biopsies were extracted after a bead-beating step with the MagNA Lyser using the MagNA Pure Compact RNA Isolation kit (Roche Molecular Systems, Inc.) including a DNase treatment. Total RNA was used to construct a cDNA library with the SMARTer Stranded Total RNA-Seq Kit - Pico Input Mammalian (Takara Bio, kit v.3). A minimum of 57 million 150bp-reads per kidney biopsy sample was processed with an agnostic in-house bioinformatics pipeline as described^9^. Sequences were aligned after translation against a viral protein reference comprehensive database (RVDB-prot.V20.0)^8^.

**SARS-CoV-2 Real-Time Polymerase Chain Reaction (RT-PCR) assay on kidney biopsies**

Total RNA was extracted from nitrogen-frozen and Optimal Cutting Temperature-embedded kidney biopsies and used to perform RT-PCR on nucleocapsid (N) and RNA-dependent RNA polymerase (RdRp) genes according to the manufacturer’s instructions (ARGENE SARS-CoV-2 R-GENE, BioMérieux, France).

**Evaluation of anti-IFN auto-Abs**

We tested all available patients’ sera from the day of diagnosis for auto-antibodies neutralizing type I IFNs using a reporter luciferase assay.

The blocking activity of anti-IFN-α2 and anti-IFN-ω auto-Abs was determined with a reporter luciferase assay. Briefly, HEK293T cells (ATCC) were transfected with a plasmid containing the Firefly luciferase gene under control of the human ISRE promoter in the pGL4.45 (Promega) backbone, and a plasmid constitutively expressing Renilla luciferase for normalization (pRL-SV40). Cells were transfected using X-tremeGene9 transfection reagent (Sigma-Aldrich, ref. number 6365779001) for 24 hours. Cells in Dulbecco’s modified Eagle medium (DMEM, Thermo Fisher Scientific) supplemented with 2% fetal calf serum (FCS) and 10% healthy control or patient serum/plasma (after inactivation at 56°C, for 20 minutes) were either left unstimulated or were stimulated with IFN-α2 (Milteny Biotec, 130-108-984), IFN-ω (Merck, SRP3061), at 10ng/mL or 100pg/mL, or IFN-β (Milteny Biotech, ref. number: 130-107-888) at 10ng/mL, for 16 hours at 37°C. Each sample was tested once for each cytokine and dose. Finally, cells were lysed for 20 minutes at room temperature and luciferase levels were measured with the Dual-Luciferase® Reporter 1000 assay system (Promega, E1980), according to the manufacturer’s protocol. Luminescence intensity was measured with a VICTOR-X Multilabel Plate Reader (PerkinElmer Life Sciences, USA). Firefly luciferase activity values were normalized to Renilla luciferase activity values. These values were then normalized against the median induction level for non-neutralizing samples, and expressed as a percentage. Samples were considered neutralizing if luciferase induction was below 15% of the median values for controls tested the same day^10^.

**Statistics**

Quantitative variables were expressed as medians and ranges and compared using the nonparametric Wilcoxon-Mann-Whitney *U* test or Student *t* test. Qualitative variables were expressed as number and percentage and were compared using the Fisher exact two-tailed test. All analyses were performed using GraphPad Prism version 9.

**Study Approval**

Research ethics board approval was granted by the local Ethics Committee of the Assistance Publique Hôpitaux de Paris (Decision number: N° 2022 0503154702). Informed consent was obtained from all parents or legal guardians; patients were informed about the purpose of the study and gave their assent.

**SUPPLEMENTARY RESULTS**

**Detailed pathology findings**

Thirty-nine (81%) patients underwent a kidney biopsy, and sixteen kidney biopsies were centralized and blindly reassessed by three kidney pathologists (Supplementary Tables S1 and S3). All 23 patients with TINUs had a kidney biopsy. The indications for kidney biopsy were intrinsic acute kidney injury (AKI) of unknown origin after exclusion of pre-renal, and obstructive causes. Indeed, most patients presented with similar clinical features including sudden polyuria and polydipsia, weight loss, fatigue and AKI, and biological inflammation. The kidney biopsy was performed at the onset of AKI diagnosis.

Three pathologists centrally and blindly reassessed the 16 biopsies of the 16 patients for whom sera stored from the day of diagnosis was available. Light microscopy analysis mainly showed acute interstitial nephritis characterized by an infiltrate of predominantly mononuclear cells associated with numerous tubulitis lesions, as classically observed in TINUs (Figure 2a and b). Immunophenotyping using immunohistochemistry (IHC) on kidney biopsies revealed that the immune cells were principally CD4 T-lymphocytes and macrophages with a relative important number of plasma cells (Figure 2, c-h). Granulomas were rarely present (1/16, 6%, Figure 2i), and eosinophil infiltrates were rather frequently observed (4/16, 25%), similar to those in non-COVID-19 associated TINUs. Of interest in the present series, frequent (7/16, 44%) polymorphonuclear cell (PMN) infiltrate was seen in the kidney interstitium with one patient (Patient 1) showing an important PMN infiltrate together with numerous PMN-mediated tubulitis resembling acute pyelonephritis lesions (Supplementary Figure 2). To investigate whether these tubulo-interstitial lesions were antibody-mediated lesions, we performed immunofluorescence analysis targeting IgG which did not reveal any tubular basal membrane immunoglobulin deposits (Figure 2j). Importantly, morphological analysis also revealed that 6/16 patients had significant renal fibrosis concerning 25 to 50% of the kidney cortex (*i.e.* FIAT 2, Supplementary Figure 3). These observed scaring lesions underline the severity of this disease, especially when it concerns young patients. Of particular interest, Patient 1 underwent two additional follow-up kidney biopsies due to unfavorable kidney outcome despite steroid therapy in association with mycophenolate mofetil, 3 and 6 months respectively, after the diagnostic biopsy. While active inflammatory lesions regressed, follow-up biopsies showed progression of renal fibrosis, further emphasizing that this disease induces fibrosis with a potential for aggravation.

To determine whether this observed renal inflammation could be directly related to a specific pathogen, we performed metagenomic next-generation sequencing (mNGS) on kidney biopsy samples from 11 patients. No definite pathogens were identified in our samples except for Patient 3, in whom a single read of BK viral genomic sequences together with astrovirus sequences were detected (Supplementary Table 1). As a low number of reads may indicate a false positive result, we decided to confirm this molecular detection by viral specific PCR, which was negative for both viruses in Patient 3. Taken together, these results indicate a false positive NGS detection of human astrovirus 1 and BKv in Patient 3.

To further investigate the presence or absence of SARS-CoV-2 mRNA in kidney samples, we performed SARS-CoV-2 RT-PCR analysis to increase detection sensitivity. Of particular interest, we detected low viral loads of SARS-CoV-2 mRNA in the kidney of two patients (Patients 1 and 2) further suggesting a potential association between COVID-19 and post-infectious aTIN. Of note, Patient 1 had a symptomatic COVID-19 infection confirmed 4 months before aTIN diagnosis by a positive nasal swab PCR. Interestingly, Patient 1 also had a pseudoneutralising test at 47% (Supplementary Table 1).

To allow *in situ* viral detection, we performed IHC against the N protein of SARS-CoV-2 and did not observe any positive staining in either kidney epithelial cells or immune infiltrating cells (Supplementary Figure 4).

All in all, we did not find any infectious agent associated with these aTINs/TINUs other than SARS-CoV-2, detected with a low viral load in only 2 kidney biopsies, supporting the hypothesis of a post-infectious pathology.

**Anti-interferon antibodies**

Auto-antibodies neutralizing type I IFNs have been shown to underlie life-threatening COVID-19 pneumonia in adults with no prior severe infection^10,11.^ We therefore sought to investigate whether such anomalies could explain the severe kidney phenotype observed in the present cohort. Available samples from 6 patients at diagnosis of aTIN were tested for auto-antibodies neutralizing type I IFNs (IFN-α2, IFN-ω, as well as IFN-β) using a luciferase assay, as previously described. We found that none of the 6 aTIN patients tested had auto-antibodies neutralizing type I IFNs **(**Supplementary Figure 5).

**SUPPLEMENTARY TABLES**

| **Patient ID Sex Age at**  **KBx** | | | **IFTA** | **Tubulitis** | **Interstitial infiltrate** | **Granuloma** | **PMN** | **Eos** | **Lymphocytes (CD3)** | **Macrophages (CD68)** | **Plasmocytes (CD138)** | **NGS** | **PCR** | **anti-S IgGs** | | **anti-N IgGs** | | **PNT** |
| --- | --- | --- | --- | --- | --- | --- | --- | --- | --- | --- | --- | --- | --- | --- | --- | --- | --- | --- |
|  |  |  |  |  |  |  |  |  |  |  |  |  |  | **LIPS** | **ABBOTT** | **LIPS** | **ABBOTT** |  |
| 1 | M | - | 2 | 3 | 3 | 0 | moderate | 0 | 3 | 3 | 1 | neg | pos | - | - | + | - | 47% |
| 2 | M | - | 2 | 3 | 3 | 0 | 0 | 0 | 2 | 2 | 1 | neg | pos | - | - | + | - | 0% |
| 3 | F | - | 2 | 3 | 3 | 0 | rare | rare | 3 | 3 | 1 | pos^*^ | neg | - | - | + | - | 0% |
| 4 | M | - | 1 | 3 | 3 | 0 | 0 | rare | 3 | 1 | 1 | neg | neg | - | - | + | - | 0% |
| 5 | M | - | 1 | 2 | 3 | 1 | rare | 0 | 2 | 2 | 1 | neg | neg | - | - | + | - | 0% |
| 6 | F | - | 1 | 3 | 2 | 0 | rare | 0 | 2 | 2 | 1 | neg | neg | - | - | + | - | 0% |
| 7 | M | - | 2 | 3 | 2 | 0 | 0 | 0 | 2 | 2 | 1 | neg | neg | - | - | + | - | 0% |
| 8 | M | - | 2 | 2 | 2 | 0 | 0 | 0 | 2 | 2 | 1 | neg | neg | - | - | + | - | 0% |
| 9 | M | - | 1 | 3 | 3 | 0 | rare | rare | 3 | 3 | 1 | neg | neg | + | + | + | - | 99% |
| 10 | F | - | 1 | 2 | 2 | 0 | 0 | 0 | 2 | 2 | 2 | neg | neg | - | - | + | - | 0% |
| 11 | F | - | 2 | 3 | 3 | 0 | 0 | 0 | 2 | 2 | 2 | neg | neg | - | - | + | - | 0% |
| 12 | M | - | 0 | 2 | 3 | 0 | 0 | 0 | 3 | 2 | 1 | NA | NA | - | - | + | - | 0% |
| 13 | M | - | 0 | 3 | 3 | 0 | rare | rare | 3 | 2 | 1 | NA | NA | - | - | + | - | 0% |
| 14 | F | - | 0 | 1 | 2 | 0 | 0 | 0 | 3 | 2 | 1 | NA | NA | - | - | + | - | 0% |
| 15 | F | - | 0 | 3 | 3 | 0 | 0 | 0 | 3 | 2 | 1 | NA | NA | - | - | + | - | 0% |
| 16 | F | - | 1 | 2 | 3 | 0 | rare | 0 | 3 | 2 | 1 | NA | NA | - | - | + | - | 0% |
| **Mean [min, max]** |  | **14.2 [9-17]** | **1.1 [0-2]** | **2.6 [1-3]** | **2.7 [2-3]** |  |  |  | **2.6 [2-3]** | **2.1 [1-3]** | **1.1 [1-2]** |  |  |  | |  | |  |
| **Ratio (%)** |  |  |  |  |  | **6.3%** | **43.8%** | **25%** |  |  |  |  |  |  | |  | |  |

**Supplementary Table 1. Histological, immunohistochemical and virological findings in COVID-19-associated aTIN/TINUs patients**

Abbreviations: IFTA: Interstitial Fibrosis and Tubular Atrophy; KBx : kidney biopsy ; NA: Not available; NGS: Next Generation Sequencing; PCR: Polymerase Chain Reaction; PMN: Polymorphonuclear cells; Eos: Eosinophils; PNT: Pseudoneutralization test; pos*: BK virus and Astrovirus

**Supplementary Table 2. Clinical and biological characteristics of the 48 children of the present cohort compared to the historical nationwide French cohort of children diagnosed with TINUs from 2000-2018**^12^

|  | **TINUs 2000-2018 (N = 46)** | | **TINUs 2020-2021 (N = 23)** | | | **aTIN/TINUs 2020-2021 (N = 48)** | | |
| --- | --- | --- | --- | --- | --- | --- | --- | --- |
|  | **n/N (%)** | median (range) | **n/N (%)** | median (range) | *p* | **n/N (%)** | median (range) | *p* |
| Female/male (sex ratio) | 24/22 (1.1 :1) |  | 13/10 (1.3 :1) |  | *0.59* | 29/19 (1.5 :1) |  | *0.53* |
| Age (years) |  | 13.8 (7.2-17) |  | 14.3 (9.6-17.6) | *0.07* |  | 14.7 (9.4-17.6) | ***0.001*** |
| **At diagnosis:** |  |  |  |  |  |  |  |  |
| **Symptoms** |  |  |  |  |  |  |  |  |
| Weight loss | 33/44 (75%) |  | 15/23 (65%) |  | *0.77* | 33/48 (69%) |  | *0.64* |
| Fever | 22/46 (48%) |  | 7/23 (30%) |  | *0.3* | 14/48 (29%) |  | *0.09* |
| Abdominal pain | 20/46 (43%) |  | 8/23 (35%) |  | *0.79* | 28/48 (58%) |  | *0.22* |
| Vomiting or diarrhea | 17/46 (37%) |  | 5/23 (22%) |  | *0.4* | 11/48 (23%) |  | *0.18* |
| Polyuro-polydipsic syndrome | 17/46 (39%) |  | 4/23 (17%) |  | *0.16* | 14/48 (29%) |  | *0.38* |
| Dyspnea | 11/46 (24%) |  | 0/23 (0%) |  | ***0.01*** | 2/48 (4%) |  | ***0.007*** |
| Arthralgia/myalgia | 9/46 (20%) |  | 1/23 (4%) |  | *0.15* | 1/48 (2%) |  | ***0.007*** |
| **Biological findings** |  |  |  |  |  |  |  |  |
| Serum creatinine (µmol/L) |  | 188.5 (82-1110) |  | 167 (63-735) | *0.89* |  | 194 (63-735) | *0.53* |
| eGFR (mL/min/1.73m²) |  | 30.6 (4.9-62.8) |  | 36.9 (8.4-87.8) | *0.7* |  | 31.9 (8.4-87.8) | *0.74* |
| eGFR > 30 mL/min/1.73m² | 24/46 (52%) |  | 13/23 (57%) |  | *0.59* | 25/48 (52%) |  | *1* |
| eGFR < 30 mL/min/1.73m² | 22/46 (48%) |  | 8/23 (35%) |  | *0.59* | 23/48 (48%) |  | *1* |
| Inflammatory syndrome |  |  |  |  |  |  |  |  |
| High CRP (mg/L) | 30/44 (68%) | 23 (0-161) | 16/22 (73%) | 21 (0-183) | *0.80* | 35/43 (81%) | 54 (0-235) | *0.16* |
| High ESR (mm/h) | 24/32 (75%) | 75.5 (22-128) | 9/12 (75%) | 61 (5-140) | *0.36* | 17/21 (81%) | 61 (5-140) | *0.35* |
| High fibrinogen (g/l) |  |  | 9/11 (81%) | 5 (2,5-9,6) |  | 19/21 (90%) | 5 (2,5-9,6) |  |
| High ferritin (µg/l) |  |  | 15/18 (83%) | 277,5 (160-740) |  | 10/12 (83%) | 272,5 (160-740) |  |
| Hyper IgG (IgG g/L) | 8/22 (36%) | 16 (8.1-27.9) | 2/17 (12%) | 13.3 (8.6-24.7) | *0.15/0.44* | 9/35 (26%) | 13.3 (6.5-25.9) | *0.55/0.28* |
| Anemia (Hb g/dL) | 35/46 (76%) | 10.5 (6.9-14.5) | 14/22 (64%) | 10.4 (7.8-14.4) | *0.76/0.91* | 22/43 (51%) | 11.2 (7.8-15.4) | *0.02/0.06* |
| Tubulopathy | 45/46 (98%) |  | 17/22 (77%) |  | *0.08* | 35/43 (81%) |  | ***0.01*** |
| Hypokalemia (mmol/L) | 10/45 (22%) | 3.8 (2.7-4.6) | 4/18 (22%) | 3.8 (3-4.4) | *0.74/0.50* | 11/38 (29%) | 3.7 (2.7-4.6) | *0.46/0.31* |
| Hypophosphatemia (mmol/L) | 10/43 (23%) | 1.13 (0.64-1.75) | 2/18 (11%) | 1.04 (0.75-1.32) | *0.48/0.09* | 4/36 (11%) | 1.03 (0.60-2.08) | *0.24/0.06* |
| Metabolic acidosis (serum bicarbonate mmol/L) | 25/45 (56%) | 20.5 (12-28) | 9/19 (47%) | 19 (12-26) | *1/0.45* | 19/38 (50%) | 19.3 (12-26) | *0.66/0.74* |
| Increased β2microglobulin (mg/L) | 28/28 (100%) |  | 13/13 (100%) |  |  | 19/22 (86%) |  | *0.08* |
| Proteinuria (mg/mmol) | 41/46 (98%) | 93.4 (8-279) | 15/22 (68%) | 59 (10-1000) | *0.16/0.07* | 34/43 (79%) | 60 (6-1000) | *0.25/0.14* |
| Normoglycemic glycosuria | 34/38 (89%) |  | 14/17 (82%) |  | *1* | 25/31 (81%) |  | *0.33* |
| Biopsy performed | 44/46 (96%) |  | 19/23 (83%) |  | *0.58* | 39/48 (81%) |  | *0.051* |
| Granuloma on kidney biopsy | 8/44 (18%) |  | 3/23 (13%) |  | *1* | 5/39 (13%) |  | *0.56* |
| Presence of uveitis | 46/46 (100%) |  | 23/23 (100%) |  |  | 23/48 (48%) |  |  |
| Occurrence of uveitis after aTIN (months) |  | 0.4 (-4 - +17) |  | 0 (-3 - +12) | *0.33* |  |  |  |
| **During follow-up:** |  |  |  |  |  |  |  |  |
| Corticotherapy | 44/46 (96%) |  | 20/23 (87%) |  | *1* | 40/48 (83%) |  | *0.09* |
| Duration of corticotherapy (months) |  | 7.3 (1.9-62) |  | 3 (1.5-10.8) | ***<0.0001*** |  | 3 (1-10.8) | ***<0.0001*** |
| Methylprednisolone pulses | 20/46 (43%) |  | 14/23 (61%) |  | *0.11* | 31/48 (65%) |  | *0.06* |
| Second-line therapy | 12/46 (26%) |  | 8/23 (34%) |  | *0.39* | 11/48 (23%) |  | *0.81* |
| Serum creatinine at last follow-up (µmol/L) |  | 70 (35-106) |  | 69 (52-154) | *0.85* |  | 73 (52-259) | *0.13* |
| eGFR at last follow-up (mL/min/1.73m²) |  | 87.5 (60.3-152.8) |  | 86 (41-117.2) | *0.68* |  | 86.0 (66.8-134.5) | *0.08* |
| **Duration of follow-up (years)** |  | **2.8 (1.0 -9.6)** |  | **1.5 (0.8-2.4)** | ***0.04*** |  | **1.4 (0.6-2.1)** | ***0.04*** |

The present cohort aTIN/TINUs 2020-2021 (N = 48) and its sub-cohort TINUs 2020-2021 (N = 23) were compared to the ‘control’ group, TINUs 2000-2018 (N = 46), a historical cohort of all consecutive children diagnosed with TINUs in France from 2000-2018^12^. Statistical comparisons were evaluated using either the Mann-Whitney U test for quantitative variables or the Fisher’s exact two-tailed test for qualitative variables. Abbreviations: aTIN: acute tubulointerstitial nephritis; CRP: C-Reactive Protein; eGFR: estimated Glomerular Filtration Rate; ESR: Erythrocyte Sedimentation Rate; Hb: Hemoglobin; TINU: tubulointerstitial nephritis and uveitis.

Out of the 48 patients, 40 received corticosteroids and 11/40 patients received a second-line immunosuppressive drug. The introduction of these treatments was not indicated by a more severe clinical, biological or histological presentation at diagnosis. Some centers include these drugs as steroid-sparing agents in patients at risk of developing corticosteroid-induced adverse events (e.g. overweight patients). Patients treated by a second-line immunosuppressive drug had similar outcomes with patients treated by steroids only at 1-year follow-up (mean eGFR of 83 ml/min/1.73m^2^ vs. 85 ml/min/1.73m^2^).

**Supplementary Table 3.** **Comparison of histological lesions in the 16 children with COVID-19-associated aTIN/TINUs and available sera, with a series of 14 children with pre-pandemic TINUs.**

|  | **TINUs - 2020-2021 (n=16)** | **TINUs - Control (n=14)** | **p** |
| --- | --- | --- | --- |
| **Sex; Female; n (%)** | 7 (44%) | 8 (57%) | 0.71 |
| **Age at KB; years** | 14.2 [9-17] | 12.9 [8-16] | 0.12 |
| **IFTA** | 1.1 [0-2] | 0.6 [0-3] | 0.09 |
| **Tubulitis** | 2.6 [1-3] | 2.1 [0-3] | 0.13 |
| **Interstitial infiltrate** | 2.7 [2-3] | 2.6 [1-3] | 0.61 |
| **Granuloma; n (%)** | 1 (6%) | 0 (0%) | 1 |
| **PMN; n (%)** | 7 (44%) | 11 (79%) | 0.07 |
| **Eos; n (%)** | 4 (25%) | 12 (86%) | <0.01 |
| **Lymphocyte (CD3)** | 2.6 [2-3] | 2.6 [1-3] | 0.73 |
| **Macrophage (CD68)** | 2.1 [1-3] | 1.7 [1-2] | 0.11 |
| **Plasmocyte (CD138)** | 1.1 [1-2] | 1.2 [1-2] | 0.65 |

Abbreviations: IFTA: Interstitial Fibrosis and Tubular Atrophy; KB: kidney biopsy; PMN: Polymorphonuclear cells; Eos: Eosinophils. Data are expressed as the mean with range [Min-Max] unless otherwise specified. Statistical comparisons were evaluated using either the Mann-Whitney U test for quantitative variables or the Fisher’s exact two-tailed test for qualitative variables.

**Supplementary Table 4. Primary antibody list.**

| **Primary antibodies** | **Supplier** | **Species** | **Type** | **Dilution** | **Final Concentration (mg/L)** | **Reference** |
| --- | --- | --- | --- | --- | --- | --- |
| CD3 | Dako | Rabbit | Polyclonal | 1/200 | 3 | GA503 |
| CD20 | Dako | Mouse | Monoclonal | 1/400 | 0.315 | GA604 |
| CD138 | Dako | Mouse | Monoclonal | 1/50 | 1.08 | M7228 |
| CD68 KP1 | Dako | Mouse | Monoclonal | 1/3000 | 0.052 | GA609 |
| CD4 | ThermoFisher | Mouse | Monoclonal | 1/100 | NA | AB_467067 |
| CD8 | Dako | Mouse | Monoclonal | 1/100 | 1.57 | GA623 |
| CD15 | Leica | Mouse | Monoclonal | 0 | NA | NCL-L-CD15-605 |
| SV40 | Roche | Mouse | Monoclonal | 0 | 0.2 | 5973775001 |
| N SARS-CoV-2 | Novusbio | Rabbit | Polyclonal | 1/500 | 0.002 | NB100-56576 |

**SUPPLEMENTARY FIGURE AND LEGENDS**

**Supplementary Figure 1. Seasonal coronaviruses seroprevalences between SARS-COV-2 N+ and N- patients**

We determined anti-N positivity of the four seasonal coronaviruses (HKU1, OC43, 229E, NL63) in our SARS-COV-2 N+ patients (n=43) compared to our SARS-COV-2 N- patients (n=26) from the Eurosurveillance study^13^. All four seasonal coronaviruses seroprevalences were comparable between SARS-COV-2 N+ and N- patients.

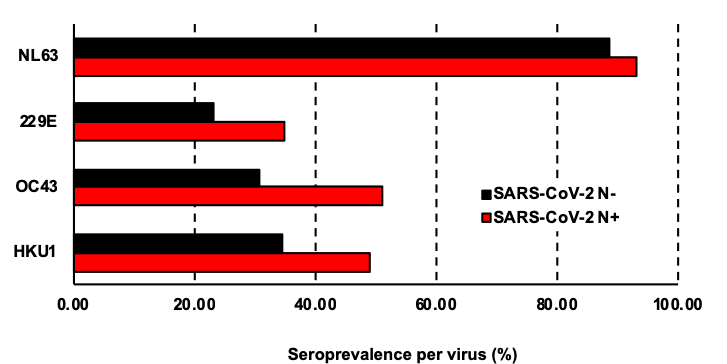

**0.31**

**0.13**

**0.66**

**0.41**

**P value**

**Supplementary Figure 2. Acute polymorphonuclear leukocytes infiltrate on patient 1 kidney biopsy.**

**a.** Light microscopy (x100) using Periodic Acid Schiff staining on patient’s 1 kidney biopsy showing diffuse interstitial infiltrate with numerous PMN leukocytes (black square). **b.** Immunohistochemistry analysis (x100) targeting CD15 on patient’s 1 kidney biopsy showing an important PMN infiltrate together with numerous tubulitis lesions.

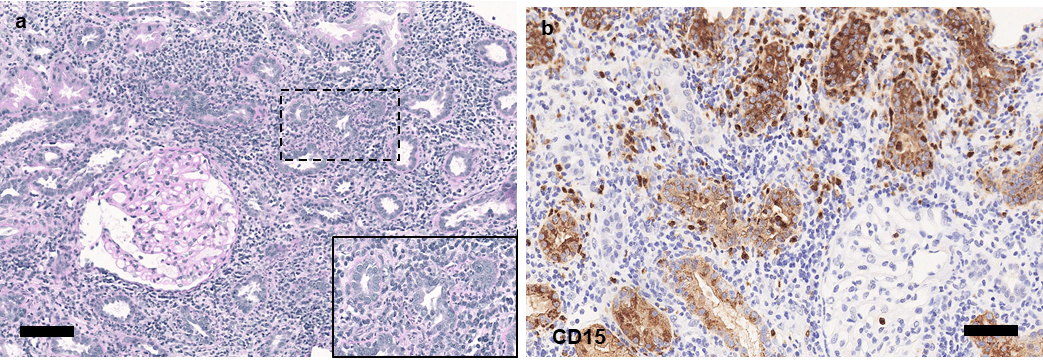

**Supplementary Figure 3. Light microscopy showing interstitial fibrosis and tubular atrophy.**

Representative images of patients with IFTA lesions. Light microscopy (x50) using Masson Trichrome staining, showing area of preserved renal parenchyma (1, Black square) and area of severe interstitial fibrosis and tubular atrophy (2, red square) with persistent interstitial infiltrate of mononuclear cells.

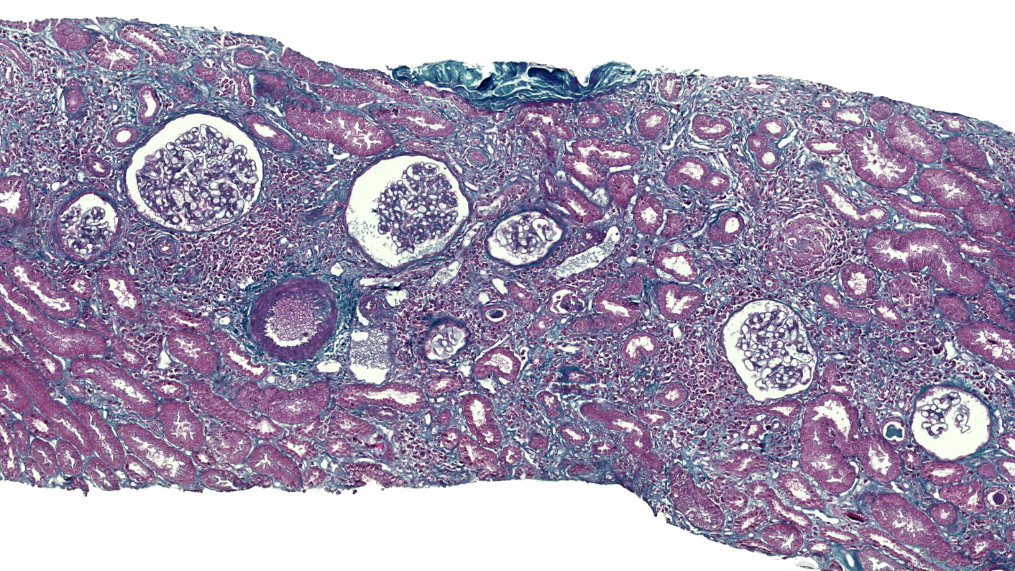

**1**

**2**

**Supplementary Figure 4. Immunohistochemical detection of N-protein of SARS-CoV-2.**

**a.** Results of SARS-CoV-2 immunostaining using an antibody directed against the nucleocapsid protein on uninfected and SARS-CoV-2 paraffin-embedded infected Vero cells. **b.** Representative image of the immunohistochemistry analysis (x100) targeting N protein of SARS-COV-2 on kidney biopsies showing no specific staining.

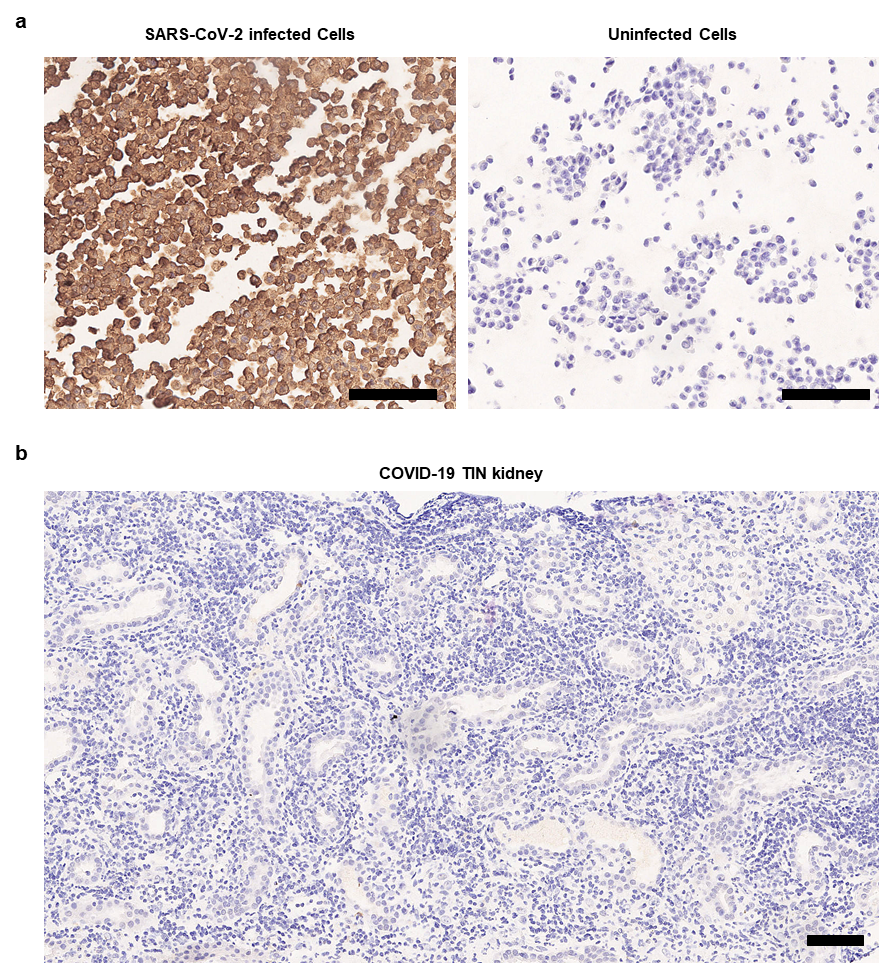

**Supplementary Figure 5. Neutralizing auto-antibodies (Abs) against IFN-α2, IFN-ω or IFN-β in children with COVID-19-related aTIN/TINUs.**

**a.** Neutralization of 10 ng/mL IFN-α2, IFN-ω or IFN-β in the presence of plasma 1/10 from pediatric patients with tubulointerstitial nephritis following COVID-19 (n=6), and one positive control (patient with neutralizing auto-Abs), suffering from APS-1 (auto-immune polyendocrine syndrome type I). Relative luciferase activity is shown (ISRE dual luciferase activity, with normalization against *Renilla* luciferase activity) after stimulation with 10ng/mL IFN-α2, IFN-ω, or IFN-β in the presence of plasma 1/10. RLA: relative luciferase activity. **b.** Neutralization of 100 pg/mL IFN-α2 or IFN-ω in the presence of plasma 1/10 from pediatric patients with tubulointerstitial nephritis following COVID-19 (n=6), and APS-1 patient.

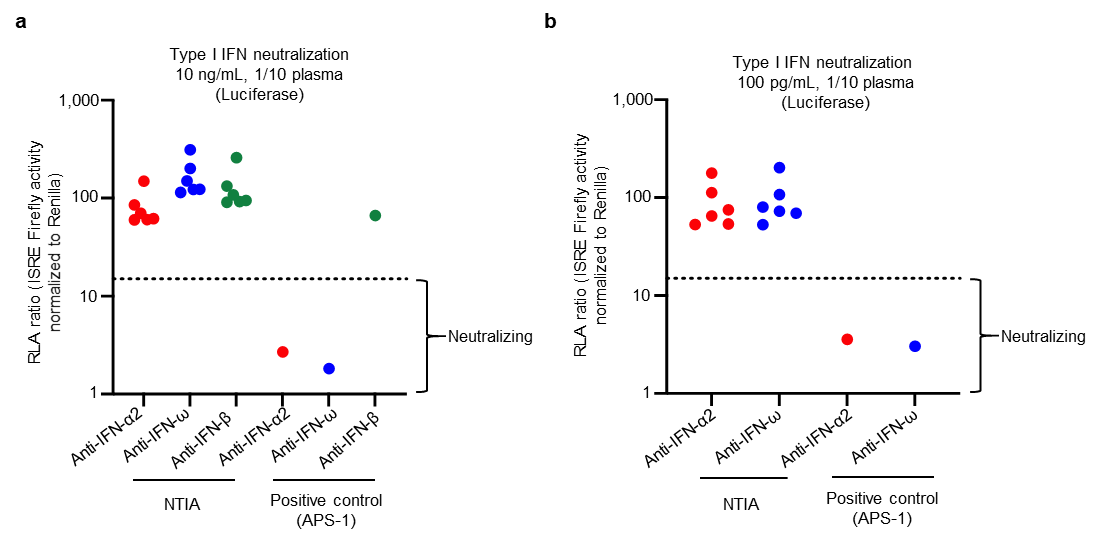
